# Prognostic association supports indexing size measures in echocardiography by body surface area

**DOI:** 10.1101/2021.12.09.21267530

**Authors:** Angus SY Fung, Dhnanjay Soundappan, Daniel E Loewenstein, David Playford, Geoffrey Strange, Rebecca Kozor, James Otton, Martin Ugander

## Abstract

**BACKGROUND:** BSA is the most commonly used metric for body size indexation of echocardiographic measures, but its use in patients who are underweight or obese is questioned (BMI<18.5 kg/m^2^ or ≥30 kg/m^2^, respectively).

**AIMS:** We aim to use survival analysis to identify an optimal body size indexation metric for echocardiographic measures that would be a better predictor of survival than body surface area (BSA) regardless of body mass index (BMI).

**METHODS:** Adult patients with no prior valve replacement were selected from the National Echocardiography Database Australia. Survival analysis was performed for echocardiographic measures both unindexed and indexed to different body size metrics, with 5-year cardiovascular mortality as the primary endpoint.

**RESULTS:** Indexation of echocardiographic measures (left ventricular end-diastolic diameter [n=230,109] and mass [n=224,244], left atrial area [n=90,596], aortic sinus diameter [n=90,805], right atrial area [n=59,516], right ventricular diameter [n=3,278], right ventricular outflow tract diameter [n=1,406]) by BSA had better prognostic performance vs unindexed measures (normal weight/overweight: average C-statistic 0.661 vs 0.620; underweight: C-statistic 0.650 vs 0.648; obese: C-statistic 0.627 vs 0.614). Indexation by other body size metrics (lean body mass, height, and/or weight raised to different powers) did not improve prognostic performance versus BSA by a clinically relevant magnitude (average C-statistic increase ≤0.02), with smaller differences in other BMI subgroups.

**CONCLUSIONS:** Indexing measures of cardiac and aortic size by BSA improves prognostic performance regardless of BMI, and no other body size metric has a clinically meaningful better performance.

## Introduction

Quantification of the dimensions of the heart and great vessels using echocardiography has both diagnostic and prognostic value for the prediction of morbidity and mortality, which also may help guide treatment in patients.^1-8^ Historically, the recommended method for body size indexation of cardiac volumes has been body surface area (BSA).^9^ More recently, recommendations for the indexation of left ventricular mass by an allometric measure of height (raised to the 2.7) have been proposed.^10, 11^ However, there is heterogeneity in the literature as to the best indexation method, and whether or not indexing of cardiac measures improves their predictive value for cardiovascular events.^6, 12-14^ In underweight and overweight patients, correction for BSA can overestimate or underestimate the prevalence of left ventricular hypertrophy, and inaccurately normalise or exaggerate indices of cardiac size.^12^ Furthermore, there are concerns regarding the physiological relationships between left ventricular mass and indexation methods. While the relationships between body surface area, height, and weight are non-linear, the indexation of left ventricular mass by these variables often assumes linear relationships.^15^ An argument can also be drawn from the theory of similarity, which reasons that relative geometries are best indexed to body size variables of similar dimensionality. For example, since left ventricular mass is related to cardiac dimensions raised to the third power, and BSA is related to a body dimension raised to the second power, it is logical that left ventricular mass should be proportional to BSA^3/2^ (also expressed as BSA^1.5^).^16^

Studies on indexation for prognostic performance have been limited in patient sample size and range of cardiac measures indexed.^5, 17-20^ Using the large-scale data available in the National Echocardiography Database of Australia (NEDA), the aim of the study was to derive one or more formulae based on height and weight to provide a method of body size indexation of cardiac and aortic measures that will be a better predictor of cardiovascular mortality than current methods based on body surface area.

## Methods

### Study design

NEDA is a large observational registry that includes routinely recorded echocardiographic data across 30 centres in Australia. Individual data linkage is used to incorporate health outcomes such as cardiovascular and all-cause mortality. The study cohort consists of patients over the age of 18 who have typically been referred clinically for imaging evaluation of known or suspected cardiovascular disease. The study was approved by the lead ethics committee at the Royal Prince Alfred Hospital (2019/ETH06989). NEDA is registered with the Australian New Zealand Clinical Trials Registry (ACTRN12617001387314). Ethical approval has been obtained from the Human Research Ethics Committees at the respective recruiting sites, and the study adheres to the Declaration of Helsinki.

### Study cohort

Echocardiographic data and basic patient characteristics were collected from participating centres from 1 January 2000 to 21 May 2019, and were transferred into a central database via an automated data extraction process. Echocardiographic measurements were made in accordance with guidelines from the American Society of Echocardiography.^9^ All data was cleaned through the removal of duplicate, inconsistent, and/or impossible measurements, and transformed into a standardized format. Individuals contributing to NEDA were assigned a unique identifier linked to their echocardiograms and their anonymity protected by stringent security protocols. As shown in Figure 1, 631,824 patients were present in the database. Of these, 182,712 (17%) patients were excluded for having less than five years of follow-up time and a further 11,282 (1%) were excluded for prior valve replacement. Echocardiograms with time from echocardiography to census or death, cause of mortality (cardiovascular and all-cause), height, weight, and the cardiac measure of interest were selected. Different populations were individually filtered for each measure of interest and analysed to maximise the number of patients for analysis. For patients with multiple echocardiograms, the earliest recorded echocardiogram was selected. Patients were grouped according to body mass index (BMI) <18.5 kg/m^2^, BMI 18.5-30 kg/m^2^, or BMI ≥30 kg/m^2^.

**Figure 1.**
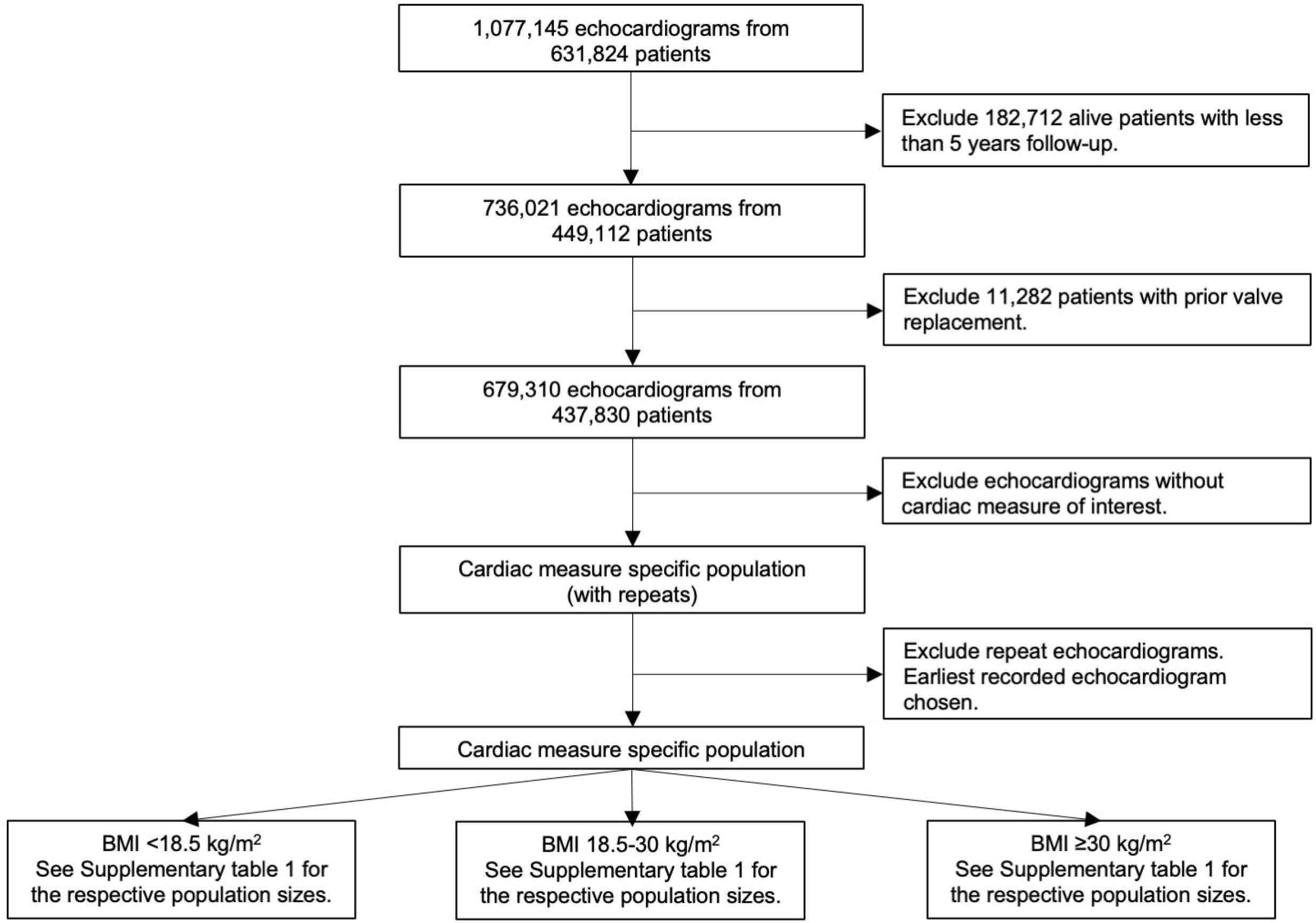
Flowchart describing patient inclusion. Exact numbers for the respective cardiac measure specific populations are given in Supplementary table 1.

### Endpoints

The primary endpoint of interest was cardiovascular mortality. Mortality data was obtained by linkage with the National Death Index.^21^ A detailed probability matching process involving patient identifiers obtained at echocardiographic recording was used to link the survival status of individuals up to the study census date of 21 May 2019. Listed causes of death were described using ICD-10 coding, which allowed for cardiovascular death to be defined (range 100-199 ICD-10AM chapter codes).^22^

### Statistical analysis

NEDA data analyses and reports were generated in agreement with STROBE guidelines.^23^ All data used in analyses were provided and no missing data was imputed. Standard procedures for describing grouped data, such as median [interquartile range (IQR)], and proportions according to patient characteristics were applied.

Cox-proportional hazard models with proportional hazards confirmed by visual inspection and numerical analysis of Schoenfeld residuals were used to derive C-statistics and hazard-ratios (with 95% confidence intervals) for the risk of cardiovascular and all-cause mortality for the entirety of the study follow-up and a five-year follow-up duration. The change in the Akaike information criterion (ΔAIC) was used to interpret the statistical robustness of the body size indexation metrics. Due to the magnification of ΔAIC by large sample sizes, the C-statistic was chosen to dictate the magnitude of difference between metrics and clinical relevance in a sample size independent fashion. Kaplan-Meier curves were constructed to visually inspect differences between indexation measures. An iterative function was coded to derive 50,000 combinations of body size metrics using different height and weight exponents according to the formula where a given body size metric = height•x weight^y. Random combinations of x and y were used as the metric for indexation for the respective echocardiographic measures for subsequent Cox-regression analysis. Cox-regression was performed using the echocardiographic measure indexed by the derived body size metric with five-year cardiovascular mortality as the endpoint to obtain the C-statistic. The C-statistic was color-coded in a scatterplot to present differences in the prognostic strength of different body size metrics including BSA as calculated according to Mosteller^24^or Du Bois^25^, lean body mass formulas by Hume^26^, Boer^27^, or James^28^, BSA raised to various powers, and height and/or weight raised to various powers. All statistical analyses were performed using R 4.0.4 (R Core Team, R Foundation for Statistical Computing, Vienna, Austria).^29^ Significance was accepted at the level of p<0.05 (two-sided).

## Results

### Study Cohort

Subject characteristics and size of the study cohorts for various cardiac and aortic measures are presented in Supplementary table 1. Due to the large sample size, differences in baseline characteristics between BMI groups were statistically significant but were not of a clinically meaningful magnitude.

### Body size metrics and mortality

Across the echocardiographic measures of right atrial area (n=59,516), right ventricular end-diastolic diameter (n=3,278), right ventricular outflow tract diameter (n=1,406), left atrial area (n=90,596), left ventricular end-diastolic diameter (n=230,109) and mass (n=224,244), and aortic sinus diameter (n=90,805), indexation by BSA as calculated by Mosteller had an average C-statistic increase from 0.620 to 0.661 for the normal weight/overweight cohort (average change in Akaike Information Criteria (ΔAIC) 708), an increase from 0.648 to 0.650 for the underweight cohort (ΔAIC 11), and an increase from 0.614 to 0.627 for the obese cohort (ΔAIC 94) compared to unindexed measures for the endpoint of five-year cardiovascular mortality as shown in Table 1.^24^ Average C-statistics are provided for composite visualisation of improvements in prognostic value across cardiac measures which followed the same trend. Indexation by other body size metrics (lean body mass, height, and/or weight raised to different powers) yielded a C-statistic increase ≤0.02. Further sex-disaggregated analysis did not differ upon visual inspection, and numerical differences between BSA and the best body size indexation metric were not clinically meaningful (average C-statistic increase ≤0.02, detailed data not shown). Smaller differences in C-statistic between indexation metrics were observed in higher BMI subgroups. Similar results (data not shown) were obtained using long-term cardiovascular mortality not limited to five years, and both long-term and five-year all-cause mortality. Furthermore, similar trends (Supplementary Figure 1-15) were observed across indexation of other aortic dimensions (sinotubular junction, ascending, root, arch) and cardiac chamber volumes (left atrial end-systolic diameter and volume, left ventricular end-diastolic volume). Across all measures, the 95% confidence interval of the C-statistic did not cross 0.50, thus the C-statistic remains significant for the normal weight/overweight group. Indexation by BSA by Mosteller provides an improvement in prognostic performance compared to the unindexed measure. Indexation metrics which have a better prognostic performance than BSA provide only a marginal improvement. However, indexation never negatively impacts prognostic performance. Kaplan-Meier curves (data not shown) did not show visually appreciable differences between indexation by BSA compared to indexation by weight across any measure.

**Table 1.** C-statistic for 5-year cardiovascular mortality both unindexed and indexed by different body size metrics for representative anatomical measures, and their average. The underweight group for LV end-diastolic diameter, RV diameter and RVOT diameter have been removed due to unreliable C-statistics stemming from small sample sizes (n=1385, n=93 and n=38, respectively). BMI = body mass index, BSA = body surface area, LBM = lean body mass, RA = right atrial, RV = right ventricular, RVOT = right ventricular outflow tract, LA = left atrial, LV = left ventricular, IVS = interventricular septum.

|  | LV mass |  |  | LA area |  |  | RA area |  |  | Aortic sinus diameter |  |  | RV diameter |  |  | LV end-diastolic diameter |  |  | RVOT diameter |  |  | Average |  |  |
| --- | --- | --- | --- | --- | --- | --- | --- | --- | --- | --- | --- | --- | --- | --- | --- | --- | --- | --- | --- | --- | --- | --- | --- | --- |
|  | BMI<br><18.5<br>kg/m<sup>2</sup> | BMI<br>18.5-30<br>kg/m<sup>2</sup> | BMI ≥30<br>kg/m<sup>2</sup> | BMI<br><18.5<br>kg/m<sup>2</sup> | BMI<br>18.5-30<br>kg/m<sup>2</sup> | BMI ≥30<br>kg/m<sup>2</sup> | BMI<br><18.5<br>kg/m<sup>2</sup> | BMI<br>18.5-30<br>kg/m<sup>2</sup> | BMI ≥30<br>kg/m<sup>2</sup> | BMI<br><18.5<br>kg/m<sup>2</sup> | BMI<br>18.5-30<br>kg/m<sup>2</sup> | BMI ≥30<br>kg/m<sup>2</sup> | BMI<br><18.5<br>kg/m<sup>2</sup> | BMI<br>18.5-30<br>kg/m<sup>2</sup> | BMI ≥30<br>kg/m<sup>2</sup> | BMI<br><18.5<br>kg/m<sup>2</sup> | BMI<br>18.5-30<br>kg/m<sup>2</sup> | BMI ≥30<br>kg/m<sup>2</sup> | BMI<br><18.5<br>kg/m<sup>2</sup> | BMI<br>18.5-30<br>kg/m<sup>2</sup> | BMI ≥30<br>kg/m<sup>2</sup> | BMI<br><18.5<br>kg/m<sup>2</sup> | BMI<br>18.5-30<br>kg/m<sup>2</sup> | BMI ≥30<br>kg/m<sup>2</sup> |
| Weight | 0.697 | 0.723 | 0.664 | 0.734 | 0.705 | 0.626 | 0.704 | 0.668 | 0.591 | 0.640 | 0.645 | 0.562 | - | 0.652 | 0.656 | - | 0.611 | 0.583 | - | 0.652 | 0.656 | 0.646 | 0.665 | 0.620 |
| Weight^1.5 | 0.686 | 0.728 | 0.660 | 0.731 | 0.697 | 0.618 | 0.708 | 0.671 | 0.589 | 0.629 | 0.634 | 0.553 | - | 0.644 | 0.643 | - | 0.609 | 0.573 | - | 0.644 | 0.643 | 0.593 | 0.661 | 0.611 |
| Height*Weight | 0.701 | 0.728 | 0.670 | 0.731 | 0.703 | 0.627 | 0.705 | 0.672 | 0.593 | 0.627 | 0.637 | 0.564 | - | 0.650 | 0.652 | - | 0.611 | 0.582 | - | 0.650 | 0.652 | 0.643 | 0.664 | 0.620 |
| BSA[Mosteller]^1.5 | 0.707 | 0.722 | 0.670 | 0.732 | 0.706 | 0.631 | 0.702 | 0.669 | 0.593 | 0.635 | 0.645 | 0.571 | - | 0.654 | 0.659 | - | 0.611 | 0.590 | - | 0.654 | 0.659 | 0.649 | 0.666 | 0.625 |
| BSA[Du Bois]^1.5 | 0.711 | 0.721 | 0.671 | 0.732 | 0.706 | 0.633 | 0.701 | 0.669 | 0.594 | 0.633 | 0.644 | 0.574 | - | 0.653 | 0.660 | - | 0.610 | 0.592 | - | 0.653 | 0.660 | 0.650 | 0.665 | 0.626 |
| BSA[Mosteller] | 0.709 | 0.710 | 0.665 | 0.731 | 0.704 | 0.632 | 0.695 | 0.661 | 0.591 | 0.642 | 0.646 | 0.576 | - | 0.650 | 0.666 | - | 0.603 | 0.593 | - | 0.650 | 0.666 | 0.650 | 0.661 | 0.627 |
| BSA[Du Bois] | 0.712 | 0.709 | 0.666 | 0.731 | 0.703 | 0.633 | 0.695 | 0.661 | 0.592 | 0.641 | 0.645 | 0.579 | - | 0.650 | 0.668 | - | 0.602 | 0.596 | - | 0.650 | 0.668 | 0.652 | 0.660 | 0.629 |
| LBM[Hume] | 0.719 | 0.715 | 0.669 | 0.726 | 0.699 | 0.627 | 0.694 | 0.663 | 0.588 | 0.624 | 0.628 | 0.564 | - | 0.644 | 0.655 | - | 0.597 | 0.582 | - | 0.644 | 0.655 | 0.653 | 0.656 | 0.620 |
| LBM[Boer] | 0.719 | 0.712 | 0.662 | 0.727 | 0.693 | 0.616 | 0.695 | 0.659 | 0.579 | 0.626 | 0.617 | 0.543 | - | 0.636 | 0.636 | - | 0.588 | 0.562 | - | 0.636 | 0.636 | 0.653 | 0.649 | 0.605 |
| LBM[James] | 0.703 | 0.714 | 0.659 | 0.726 | 0.695 | 0.615 | 0.697 | 0.660 | 0.577 | 0.626 | 0.619 | 0.546 | - | 0.637 | 0.635 | - | 0.588 | 0.562 | - | 0.637 | 0.635 | 0.643 | 0.650 | 0.604 |
| BMI | 0.678 | 0.693 | 0.637 | 0.724 | 0.690 | 0.611 | 0.687 | 0.647 | 0.579 | 0.624 | 0.615 | 0.533 | - | 0.632 | 0.646 | - | 0.581 | 0.558 | - | 0.632 | 0.646 | 0.619 | 0.642 | 0.601 |
| Height^2.7 | 0.726 | 0.706 | 0.672 | 0.725 | 0.697 | 0.635 | 0.692 | 0.662 | 0.594 | 0.620 | 0.618 | 0.580 | - | 0.642 | 0.662 | - | 0.595 | 0.596 | - | 0.642 | 0.662 | 0.665 | 0.652 | 0.629 |
| Height^2.13 | 0.724 | 0.702 | 0.669 | 0.727 | 0.698 | 0.636 | 0.690 | 0.659 | 0.593 | 0.628 | 0.621 | 0.584 | - | 0.642 | 0.666 | - | 0.593 | 0.600 | - | 0.642 | 0.666 | 0.668 | 0.651 | 0.631 |
| Height^2 | 0.724 | 0.701 | 0.668 | 0.727 | 0.697 | 0.636 | 0.690 | 0.658 | 0.593 | 0.630 | 0.621 | 0.585 | - | 0.642 | 0.667 | - | 0.592 | 0.600 | - | 0.642 | 0.667 | 0.668 | 0.650 | 0.631 |
| Height^1.5 | 0.720 | 0.695 | 0.663 | 0.727 | 0.695 | 0.635 | 0.687 | 0.653 | 0.591 | 0.635 | 0.618 | 0.585 | - | 0.640 | 0.669 | - | 0.586 | 0.599 | - | 0.640 | 0.669 | 0.667 | 0.647 | 0.630 |
| Height | 0.715 | 0.688 | 0.657 | 0.725 | 0.692 | 0.632 | 0.683 | 0.648 | 0.589 | 0.635 | 0.607 | 0.579 | - | 0.634 | 0.670 | - | 0.577 | 0.594 | - | 0.634 | 0.670 | 0.664 | 0.640 | 0.627 |
| Unindexed | 0.702 | 0.670 | 0.641 | 0.718 | 0.679 | 0.622 | 0.672 | 0.635 | 0.582 | 0.613 | 0.568 | 0.552 | - | 0.617 | 0.666 | - | 0.553 | 0.572 | - | 0.617 | 0.666 | 0.648 | 0.620 | 0.614 |

Figure 2 shows how indexation by different body size metrics (based on the formula height•x weight^y) perform prognostically, averaged across the representative cardiac and aortic anatomical measure. The color scale shows increasing C-statistics, where each color change represents one percentage point of C-statistic. Numerical values for selected measures are presented in Table 1. In summary, in the normal weight/overweight cohort, indexation by

**Figure 2.**
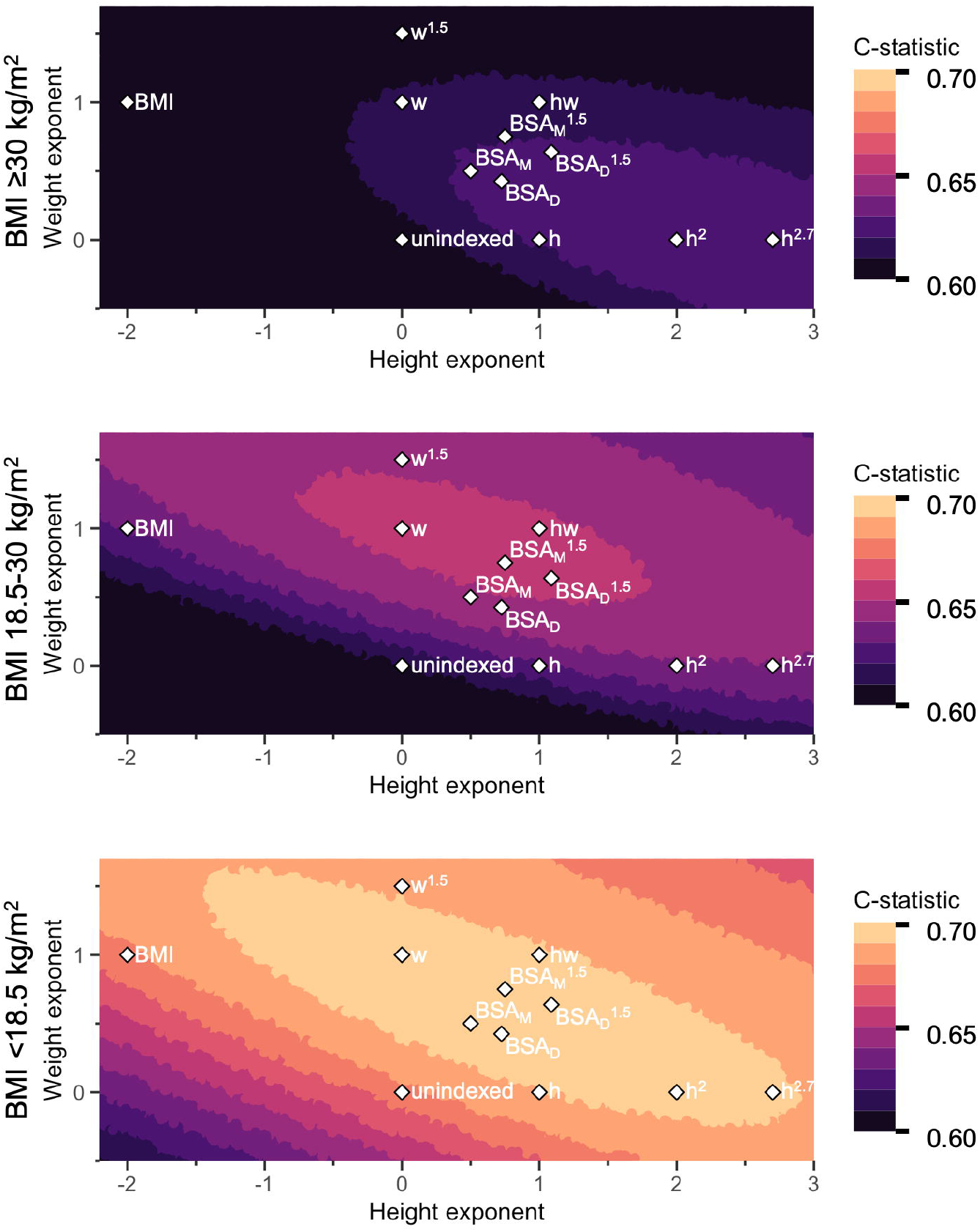
Average prognostic strength for predicting 5-year cardiovascular mortality when indexing for body size for right atrial area, right ventricular diameter, right ventricular outflow tract diameter, left atrial area, left ventricular diameter, left ventricular mass, and aortic sinus diameter. The axes represent the height and weight exponents of a body size indexation metric of the format height•x * weight^y. The color scale shows the prognostic strength from a C-statistic of 0.60 to 0.70, each increment representing a 1 %-point improvement. Existing body size metrics were plotted: h = height, w = weight, hw = height * weight, BMI = body mass index, BSA = body surface area, M = Mosteller, D = DuBois. In the normal weight/overweight group, from unindexed to body surface area by Mosteller there is a 4%-point improvement, but further improvement is limited to <1%-point. Similar trends can be observed in the underweight and obese group. See text for details.

BSA improves prognostic performance compared to unindexed measures by four percentage points of the C-statistic. Further improvement beyond BSA is limited to <1 percentage point improvement on average. Measures of left ventricular end-diastolic diameter, right ventricular end-diastolic diameter and right ventricular outflow tract diameter were not included in the underweight group due to unreliable C-statistic measure stemming from small sample sizes (n=1385, n=93 and n=38, respectively). Indexation by BSA yielded smaller improvements in prognostic performance in the underweight and obese cohort, but indexation by any other body size metric did not provide any meaningfully stronger association with survival. Further analyses in obese populations and higher BMI subgroups (BMI 30-35, 35-40 and >40) showed that BSA performed similarly to height raised to various powers (data not shown).

## Discussion

In this study of body size indexation of cardiac and aortic sizes using real-world echocardiographic data from a large-scale nationwide cohort, indexation by BSA is shown to improve prognostic performance compared to unindexed measures regardless of BMI. Furthermore, no other body size indexation metric provided any meaningful improvement in prognostic performance beyond BSA. The current study comprehensively assessed indexation metrics with varying height and weight exponents in different forms (multiplicative, additive/subtractive, both), and in sex-specific cohorts. It was not possible to derive a body size metric with clinically meaningfully better prognostic performance than BSA. This means that using mortality as the arbiter of indexation effectiveness, no other indexation method exists that is clinically meaningfully better than BSA across all investigated cardiac measures. In accordance with current guidelines for echocardiography^9^, cardiac measures can continue to be indexed using any formula for BSA, regardless of BMI or the echocardiographic measure of interest.

### Comparison with indexation by height

The current study found that indexation by BSA improves prognostic performance compared to height regardless of BMI. It has been suggested that indexation by height improves detection of left ventricular hypertrophy and associations with cardiovascular events and mortality compared to BSA in obese populations.^14, 20, 30^ Recently, it has been shown that indexation by height for left atrial volumes was able to maintain proportionality and avoid overcorrection for body size.^31, 32^ However, a consideration of prognostic value is more useful clinically than allometry. While indexation by height, height^2^ and modified versions of BSA have been shown to reclassify patients into binary groups with mortality benefit, cut-off values for these groups are arbitrary and fail to appreciate the continuous nature of such variables.^33^ Further, other studies have found no improvement in indexation by height compared to BSA.^5, 34^ The findings of the current study confirm that indexation by height does not provide additional prognostic value compared to indexation by BSA across any BMI group.

### Sex differences

The current study found that indexation of cardiac measures disaggregated by sex does not improve prognostic performance. Differences in left ventricular mass have been found between male and female patients.^35, 36^ A consideration of sex in the indexation of cardiac measures measured by cardiac magnetic resonance imaging has been suggested for improved prediction of incident heart failure.^13^ The current study analysed sex-specific cohorts and found that body size metrics derived from sex-specific cohorts were interchangeable with negligible differences in prognostic performance. Thus, the findings of the current study suggest that the relationship between cardiac or aortic size and survival in relation to body size does not fundamentally differ between the sexes. Notably, this is still consistent with using sex-specific cut-offs for normality for a given measure.

### Strengths and limitations

The limitations of applying and interpreting big data in NEDA have been acknowledged previously.^22, 37^ At the time of analysis, NEDA did not include important clinical details of common conditions such as coronary artery disease, ischaemic heart disease, and clinically diagnosed HF, which may impact mortality. Prevalent cardiovascular disease, namely myocardial infarction, can alter the natural relationship between anthropometric parameters and heart structures, and this could not be accounted for in this study.^38^ That said, the current study used large-scale, real-world data with relevant clinical outcomes that inevitably have measurement variability between centres and observers. While on one hand, this is a limitation as an uncontrolled source of data heterogeneity, it is in fact a strength that reinforces the integrity of observed trends that exist despite sources of variability.

The use of data from NEDA also results in confounding by indication, wherein the inclusion of patients is biased by their need for an echocardiogram. However, this is not a limitation but rather a strength as indexation metrics are intended to be applied to such patients.

The main endpoint of consideration, cardiovascular mortality, was linked from the Australian National Death Index which has a high sensitivity and specificity (93% and 90%, respectively) validating its use as the primary endpoint.^21^ Furthermore, in the current study, similar trends were also observed across all-cause mortality, further reinforcing the validity of the results.

The current study did not include the impact of age on aortic size or mortality which may be a source of uncontrolled confounding. An increase in age has long been established to be related to an increased aortic size, which would affect interpretation of the indexed measure.^39, 40^ The association between age and mortality is both intuitive and widely accepted.^41, 42^ However, a consideration of age in choice of body size indexation metric is impractical clinically, and fails to achieve the goal of indexation, namely, to account for body size. Given the strong association of age with mortality, the inclusion of age into statistical models overcasts differences between indexation metrics. A consideration of age is more appropriate for cut-off values of the indexed cardiac and aortic size measure, but not necessarily for the choice of body size indexation metric. Importantly, similar age distributions existed between the respective cohorts in the current study. Thus, while theoretically attractive, consideration of the effect of age does not contribute to addressing the purpose of the optimal choice of body size indexation method.

The NEDA cohort comprises patients being investigated for known or suspected cardiovascular disease. Data was largely obtained from specialist centres or clinics in Australia. While the NEDA cohort is representative of the diverse and multiethnic population in Australia with a largely high-functioning level of health care, applicability may be different in other contexts.

The current study has definitively demonstrated that measures of cardiac and aortic size should continue to be indexed by BSA regardless of BMI. No other existing or derived body size metric (lean body mass or height and/or weight raised to various powers) is clinically meaningfully better. Left atrial size and left ventricular mass indexed to BSA provided the strongest prognostic association of all transthoracic echocardiographic size measures.

## Data Availability

All data produced in the present work are contained in the manuscript (unless otherwise specified). Supplementary data in the present study is available upon reasonable request to the authors.

## Acknowledgements

The authors gratefully acknowledge the NEDA organisation and it contributing sites and investigators for the creation of the NEDA database.

