## Supplementary Table 1 for "Prognostic association supports indexing size measures in echocardiography by body surface area"

**Supplementary table 1.** Subject characteristics for patients with available follow-up data for the respective cardiac or aortic measures. RA = right atrial, RV = right ventricular, RVOT = right ventricular outflow tract, LA = left atrial, LV = left ventricular, IVS = interventricular septum.

| | **RA area** | | | | **RV diameter** | | | | **RVOT diameter** | | | |
|---|---|---|---|---|---|---|---|---|---|---|---|---|
| | **BMI <18.5 kg/m$^2$** | **BMI 18.5-30 kg/m$^2$** | **BMI ≥30 kg/m$^2$** | **All** | **BMI <18.5 kg/m$^2$** | **BMI 18.5-30 kg/m$^2$** | **BMI ≥30 kg/m$^2$** | **All** | **BMI <18.5 kg/m$^2$** | **BMI 18.5-30 kg/m$^2$** | **BMI ≥30 kg/m$^2$** | **All** |
| Subjects, n (%) | 1413 (2) | 38280 (64) | 19823 (33) | 59516 (100) | 93 (3) | 2210 (67) | 975 (30) | 3278 (100) | 38 (3) | 995 (71) | 373 (27) | 1406 (100) |
| Events, n (%) | 186 (13.2) | 3137 (8.2) | 1290 (6.5) | 4613 (7.8) | 15 (16.1) | 211 (9.5) | 82 (8.4) | 308 (9.4) | 6 (15.8) | 111 (11.2) | 34 (9.1) | 151 (10.7) |
| Female sex, n (%) | 1012 (72) | 17964 (47) | 9767 (49) | 28743 (48) | 46 (50) | 751 (34) | 374 (38) | 1171 (36) | 26 (68) | 470 (47) | 189 (51) | 685 (49) |
| Age, years | 68 [39-81] | 66 [51-77] | 64 [52-73] | 65 [52-76] | 60 [39-79] | 63 [46-75] | 60 [48-69] | 62 [47-74] | 50 [26-70] | 56 [36-75] | 61 [45-72] | 57 [38-74] |
| Follow-up, years | 5.6 [2.0-8.2] | 7.2 [5.1-11.2] | 7.2 [5.3-10.6] | 7.1 [5.2-11] | 6.6 [3.2-9.4] | 7 [4.7-9.6] | 7.2 [5.5-9.7] | 7 [5.1-9.6] | 5.8 [2.5-7] | 6.1 [3.7-7.5] | 6 [5.0-7.6] | 6 [4.1-7.5] |
| | **LA area** | | | | **LA diameter** | | | | **LA volume** | | | |
| | **BMI <18.5 kg/m$^2$** | **BMI 18.5-30 kg/m$^2$** | **BMI ≥30 kg/m$^2$** | **All** | **BMI <18.5 kg/m$^2$** | **BMI 18.5-30 kg/m$^2$** | **BMI ≥30 kg/m$^2$** | **All** | **BMI <18.5 kg/m$^2$** | **BMI 18.5-30 kg/m$^2$** | **BMI ≥30 kg/m$^2$** | **All** |
| Subjects, n (%) | 2259 (2) | 58597 (65) | 29740 (33) | 90596 (100) | 2315 (3) | 59425 (66) | 27713 (31) | 89453 (100) | 3314 (2) | 101920 (68) | 45306 (30) | 150540 (100) |
| Events, n (%) | 302 (13.4) | 4901 (8.4) | 1996 (6.7) | 7199 (7.9) | 227 (9.8) | 4137 (7.0) | 1542 (5.6) | 5906 (6.6) | 455 (13.7) | 6924 (6.8) | 2214 (4.9) | 9593 (6.4) |
| Female sex, n (%) | 1573 (70) | 26390 (45) | 14269 (48) | 42232 (47) | 1572 (68) | 28680 (48) | 14384 (52) | 44636 (50) | 2410 (73) | 47765 (47) | 22504 (50) | 72679 (48) |
| Age, years | 67 [39-81] | 66 [51-77] | 63 [52-72] | 65 [51-76] | 63 [35-78] | 63 [47-75] | 62 [51-71] | 63 [49-74] | 71 [45-83] | 65 [51-77] | 62 [52-72] | 64 [51-76] |
| Follow-up, years | 5.4 [1.7-9] | 7.4 [5-10.8] | 7.6 [5.3-10.5] | 7.4 [5.1-10.6] | 6.6 [2.6-11] | 8.4 [5.4-12.7] | 8.2 [5.6-11.8] | 8.3 [5.5-12.4] | 5.7 [2-8.8] | 7.6 [5.4-10.3] | 7.9 [5.7-10.4] | 7.7 [5.5-10.3] |
| | **LV end-diastolic diameter** | | | | **LV end-diastolic volume** | | | | **IVS thickness** | | | |
| | **BMI <18.5 kg/m$^2$** | **BMI 18.5-30 kg/m$^2$** | **BMI ≥30 kg/m$^2$** | **All** | **BMI <18.5 kg/m$^2$** | **BMI 18.5-30 kg/m$^2$** | **BMI ≥30 kg/m$^2$** | **All** | **BMI <18.5 kg/m$^2$** | **BMI 18.5-30 kg/m$^2$** | **BMI ≥30 kg/m$^2$** | **All** |
| Subjects, n (%) | 6375 (3) | 155211 (67) | 68523 (30) | 230109 (100) | 1385 (2) | 46330 (72) | 16305 (25) | 64020 (100) | 6468 (3) | 157507 (68) | 69270 (30) | 233245 (100) |
| Events, n (%) | 673 (10.6) | 10938 (7.0) | 3684 (5.4) | 15295 (6.6) | 177 (12.8) | 2889 (6.2) | 668 (4.1) | 3734 (5.8) | 677 (10.5) | 11160 (7.1) | 3722 (5.4) | 15559 (6.7) |
| Female sex, n (%) | 4287 (67) | 72233 (47) | 34031 (50) | 110551 (48) | 1037 (75) | 21691 (47) | 8268 (51) | 30996 (48) | 4361 (67) | 73126 (46) | 34398 (50) | 111885 (48) |
| Age, years | 66 [42-80] | 64 [49-76] | 62 [51-71] | 63 [50-75] | 73 [47-84] | 65 [50-77] | 62 [51-72] | 64 [50-76] | 66 [42-80] | 64 [49-76] | 62 [51-71] | 63 [50-75] |
| Follow-up, years | 6.1 [2.1-9.8] | 7.3 [5.2-10.6] | 7.4 [5.5-10.4] | 7.3 [5.3-10.5] | 5.6 [2.1-8.2] | 7 [5.4-9.3] | 7.2 [5.6-9.3] | 7 [5.4-9.3] | 6.1 [2.1-9.8] | 7.3 [5.2-10.5] | 7.4 [5.5-10.3] | 7.3 [5.3-10.4] |

RA = right atrial, RV = right ventricular, RVOT = right ventricular outflow tract, LA = left atrial, LV = left ventricular, IVS = interventricular septum.

**Supplementary table 1.** Subject characteristics for patients with available follow-up data for the respective cardiac or aortic measures. RA = right atrial, RV = right ventricular, RVOT = right ventricular outflow tract, LA = left atrial, LV = left ventricular, IVS = interventricular septum.

| | **LV mass** | | | | **Aortic sinus diameter** | | | | **Aorta at sinotubular diameter** | | | |
|---|---|---|---|---|---|---|---|---|---|---|---|---|
| | **BMI <18.5 kg/m²** | **BMI 18.5-30 kg/m²** | **BMI ≥30 kg/m²** | **All** | **BMI <18.5 kg/m²** | **BMI 18.5-30 kg/m²** | **BMI ≥30 kg/m²** | **All** | **BMI <18.5 kg/m²** | **BMI 18.5-30 kg/m²** | **BMI ≥30 kg/m²** | **All** |
| Subjects, n (%) | 6229 (3) | 151563 (68) | 66452 (30) | 224244 (100) | 2157 (2) | 62533 (69) | 26115 (29) | 90805 (100) | 1334 (2) | 39876 (66) | 19403 (32) | 60613 (100) |
| Events, n (%) | 650 (10.4) | 10583 (7.0) | 3521 (5.3) | 14754 (6.6) | 284 (13.2) | 4237 (6.8) | 1244 (4.8) | 5765 (6.3) | 199 (14.9) | 3189 (8.0) | 1128 (5.8) | 4516 (7.5) |
| Female sex, n (%) | 4205 (68) | 70812 (47) | 33210 (50) | 108227 (48) | 1551 (72) | 28663 (46) | 12599 (48) | 42813 (47) | 922 (69) | 16624 (42) | 8673 (45) | 26219 (43) |
| Age, years | 66 [42-80] | 64 [49-76] | 62 [51-71] | 63 [50-75] | 72 [45-83] | 65 [51-78] | 62 [52-72] | 64 [51-76] | 72 [45-83] | 68 [54-78] | 64 [54-73] | 67 [54-77] |
| Follow-up, years | 6.1 [2.1-9.9] | 7.3 [5.2-10.6] | 7.4 [5.5-10.3] | 7.3 [5.3-10.5] | 5.7 [1.8-8.4] | 7.5 [5.4-9.7] | 7.6 [5.7-9.6] | 7.5 [5.4-9.7] | 5.5 [1.6-8.6] | 7.7 [5.2-10.6] | 7.8 [5.7-10.5] | 7.7 [5.3-10.5] |
| | **Aortic root diameter** | | | | **Ascending aorta diameter** | | | | **Aortic arch diameter** | | | |
| | **BMI <18.5 kg/m²** | **BMI 18.5-30 kg/m²** | **BMI ≥30 kg/m²** | **All** | **BMI <18.5 kg/m²** | **BMI 18.5-30 kg/m²** | **BMI ≥30 kg/m²** | **All** | **BMI <18.5 kg/m²** | **BMI 18.5-30 kg/m²** | **BMI ≥30 kg/m²** | **All** |
| Subjects, n (%) | 4773 (2) | 133153 (68) | 57212 (29) | 195138 (100) | 2693 (3) | 65077 (64) | 33619 (33) | 101389 (100) | 454 (3) | 10988 (65) | 5405 (32) | 16847 (100) |
| Events, n (%) | 541 (11.3) | 8971 (6.7) | 2844 (5.0) | 12356 (6.3) | 295 (11.0) | 5587 (8.6) | 2252 (6.7) | 8134 (8.0) | 43 (9.5) | 803 (7.3) | 312 (5.8) | 1158 (6.9) |
| Female sex, n (%) | 3272 (69) | 62640 (47) | 28806 (50) | 94718 (49) | 1770 (66) | 28766 (44) | 15877 (47) | 46413 (46) | 303 (67) | 4667 (43) | 2393 (44) | 7363 (44) |
| Age, years | 68 [41-81] | 64 [49-76] | 62 [51-71] | 63 [50-75] | 66 [43-80] | 67 [52-78] | 63 [53-73] | 65 [52-76] | 52 [26-77] | 64 [47-76] | 61 [50-70] | 63 [48-74] |
| Follow-up, years | 5.9 [2.1-9.6] | 7.6 [5.4-11] | 7.7 [5.6-10.7] | 7.6 [5.4-10.9] | 5.9 [1.9-9.6] | 6.7 [4.5-9.6] | 6.9 [5.2-9.6] | 6.8 [4.9-9.6] | 5.8 [2.1-8.7] | 6.6 [5.2-9] | 6.8 [5.3-8.9] | 6.7 [5.2-9] |

RA = right atrial, RV = right ventricular, RVOT = right ventricular outflow tract, LA = left atrial, LV = left ventricular, IVS = interventricular septum.
