## Supplementary Figure 1-15 for "Prognostic association supports indexing size measures in echocardiography by body surface area"

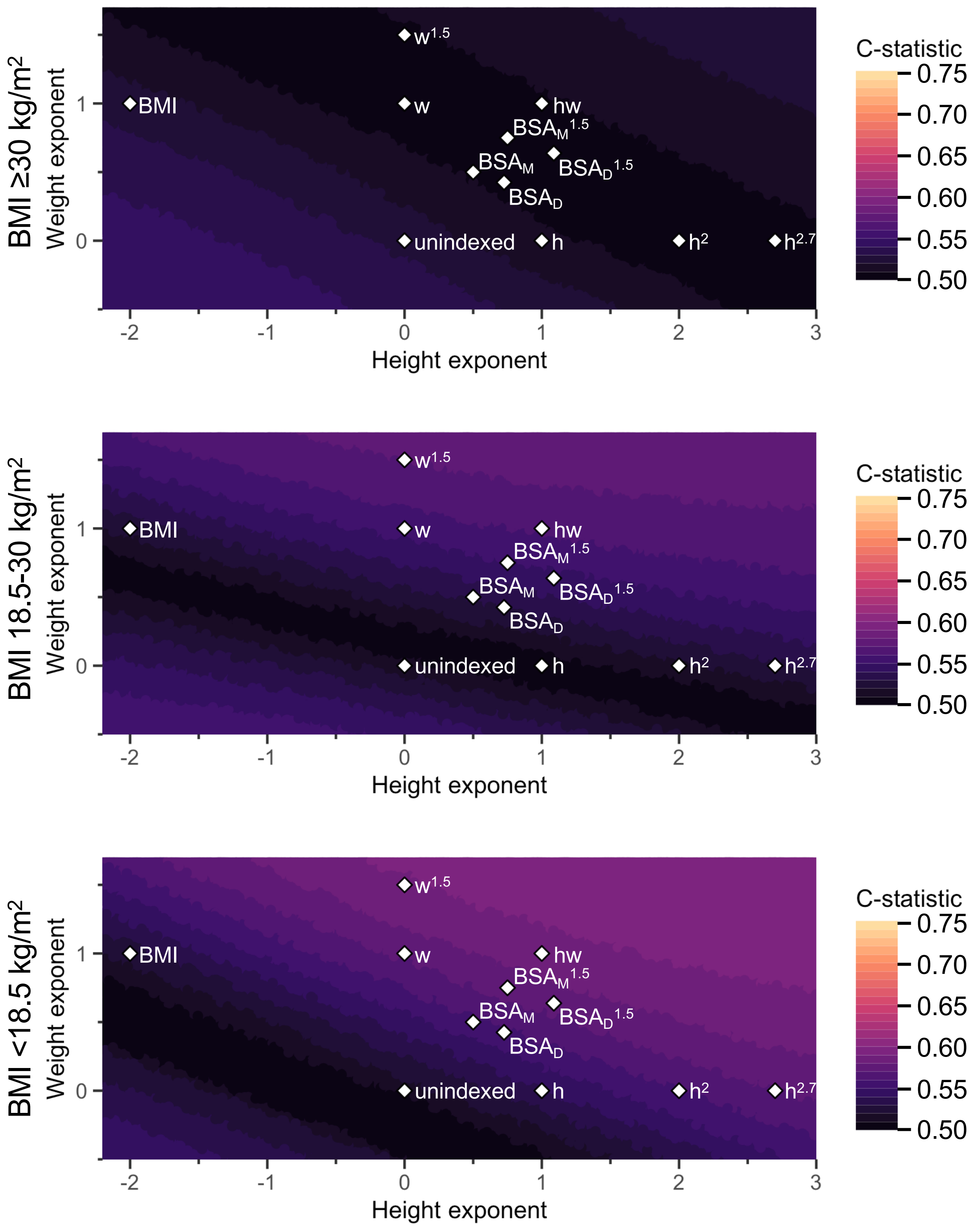


**Supplemental Figure 1.** Average prognostic strength of indexing for body size in aorta at sinotubular diameter.

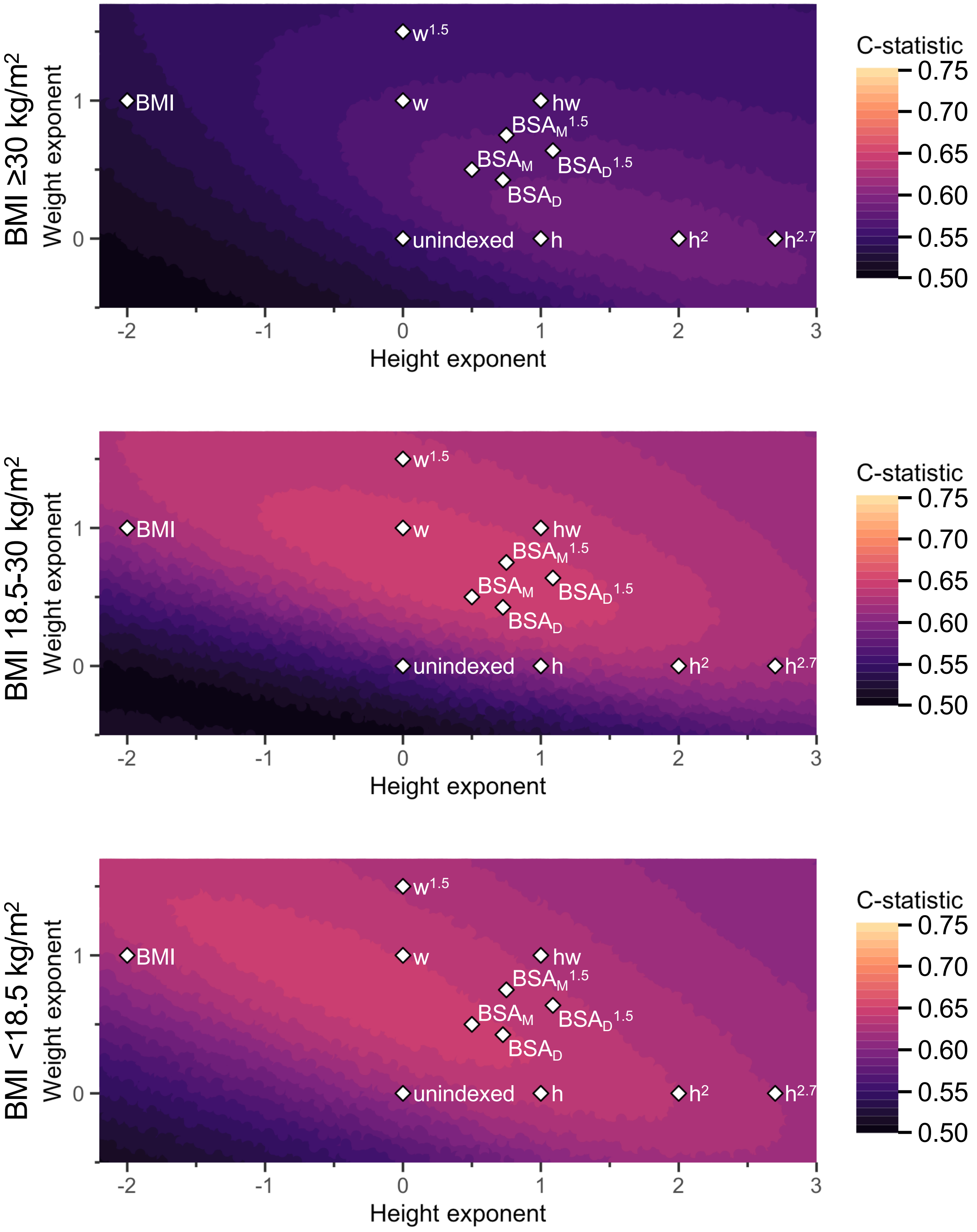


**Supplemental Figure 2.** Average prognostic strength of indexing for body size in aortic sinus diameter.

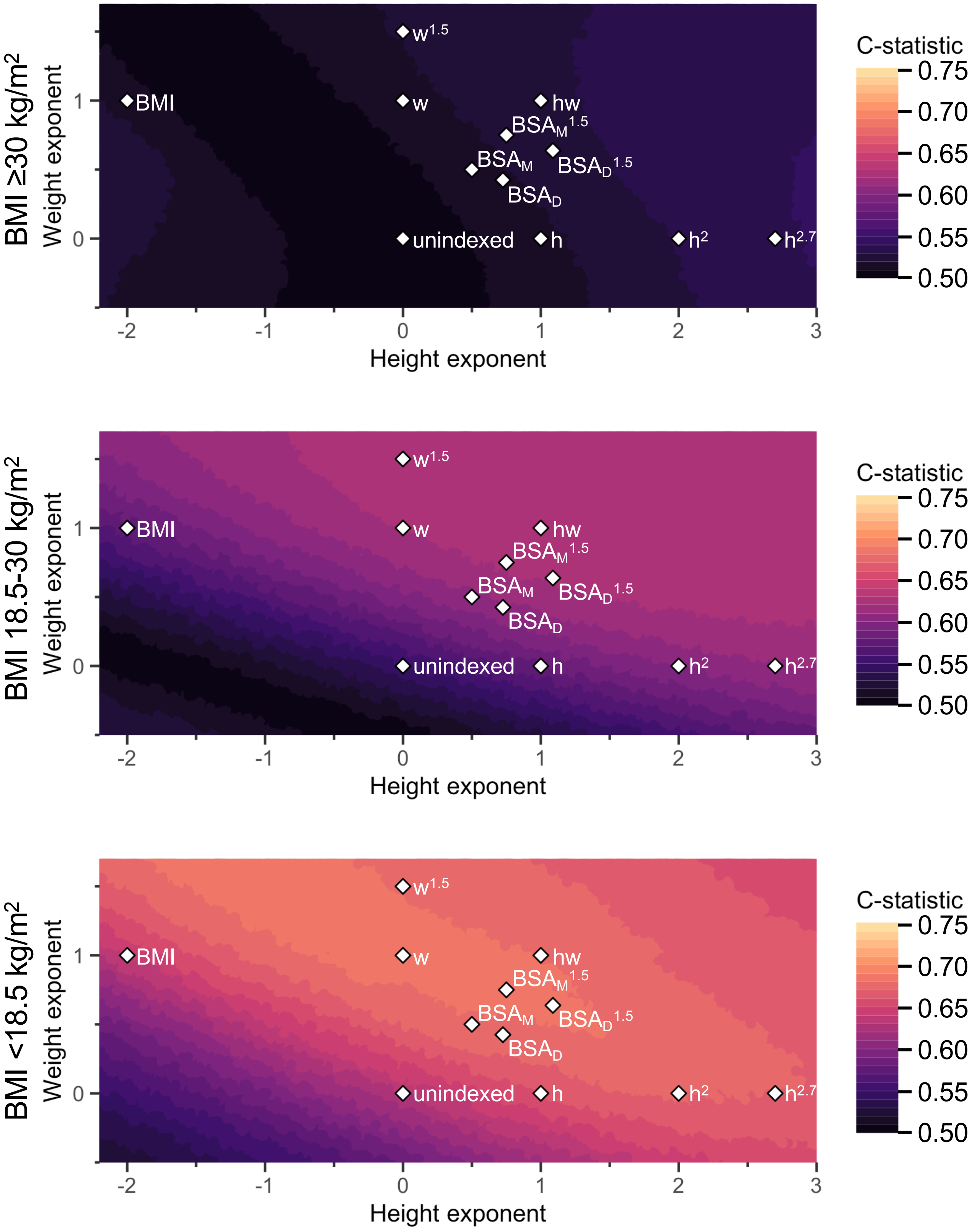


**Supplemental Figure 3.** Average prognostic strength of indexing for body size in aortic arch diameter.

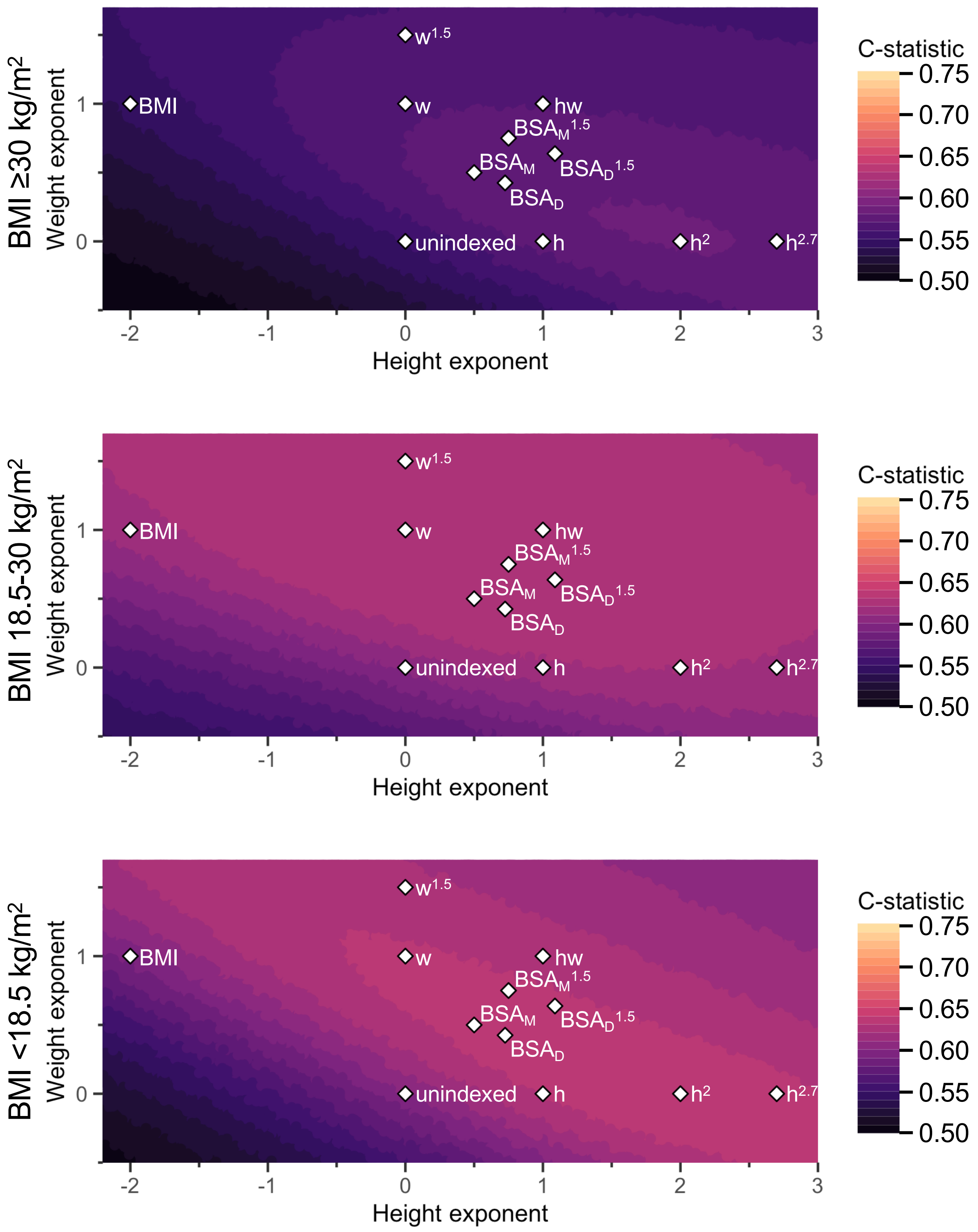


**Supplemental Figure 4.** Average prognostic strength of indexing for body size in aortic root diameter.

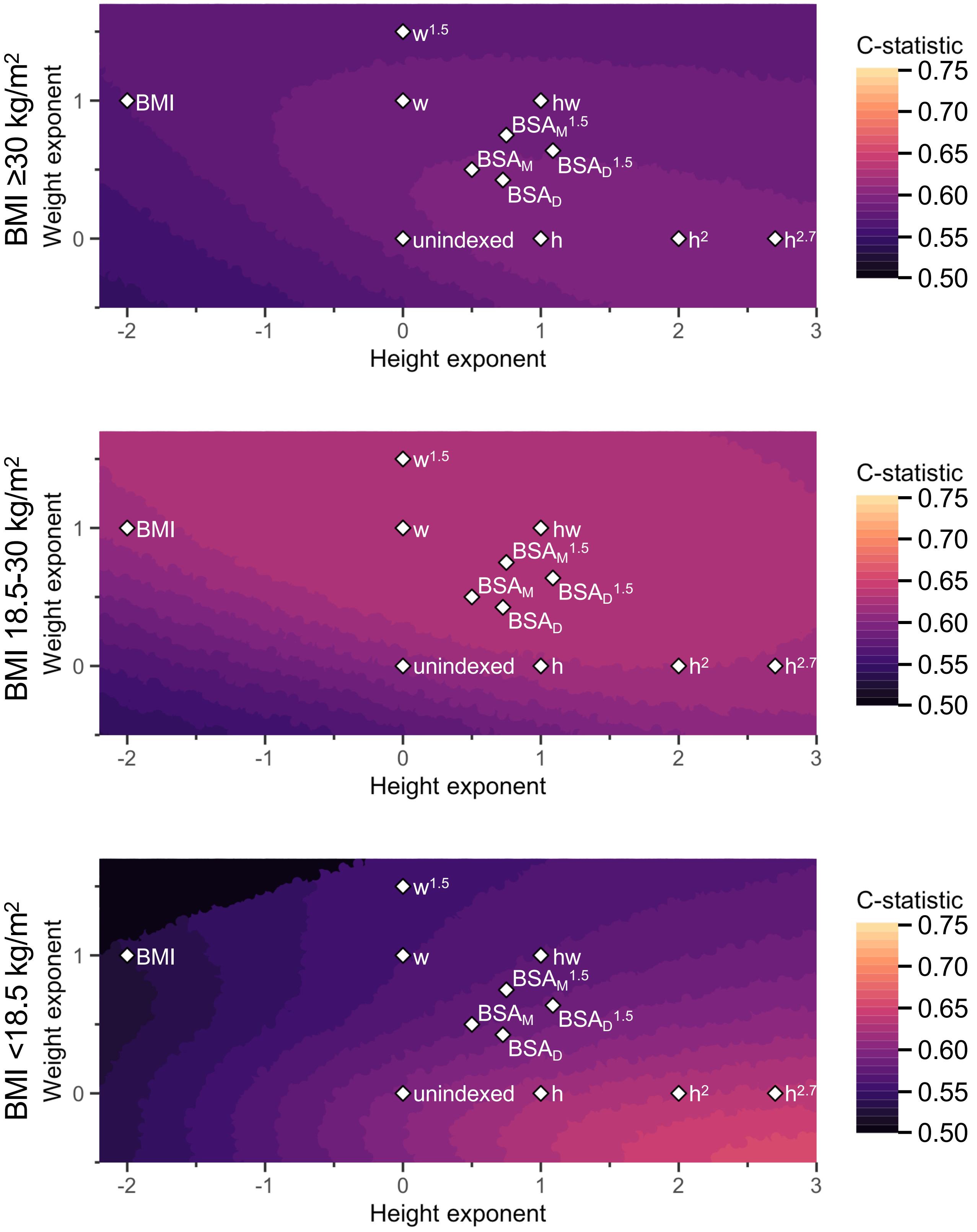


**Supplemental Figure 5.** Average prognostic strength of indexing for body size in ascending aorta diameter.

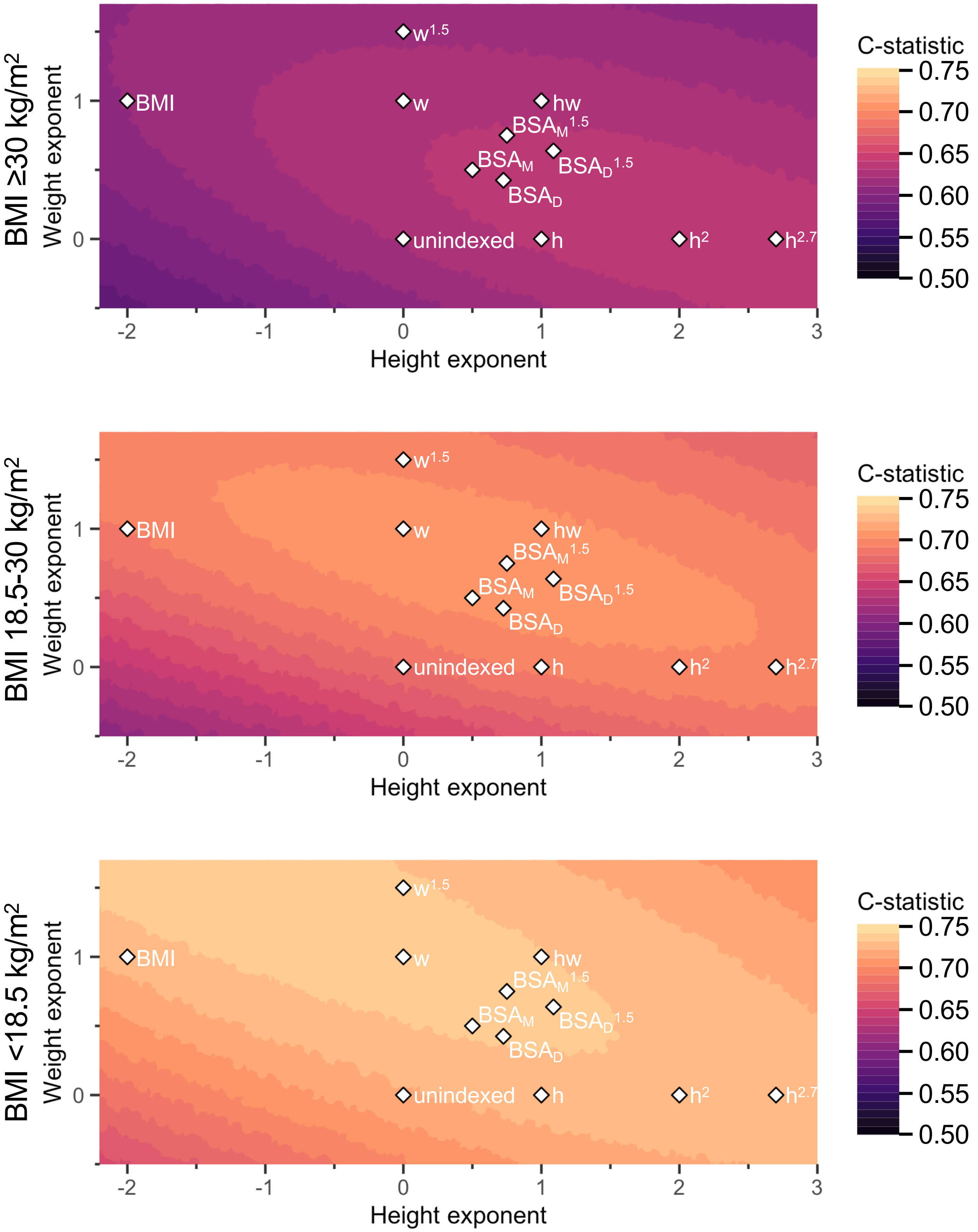


**Supplemental Figure 6.** Average prognostic strength of indexing for body size in LA area.

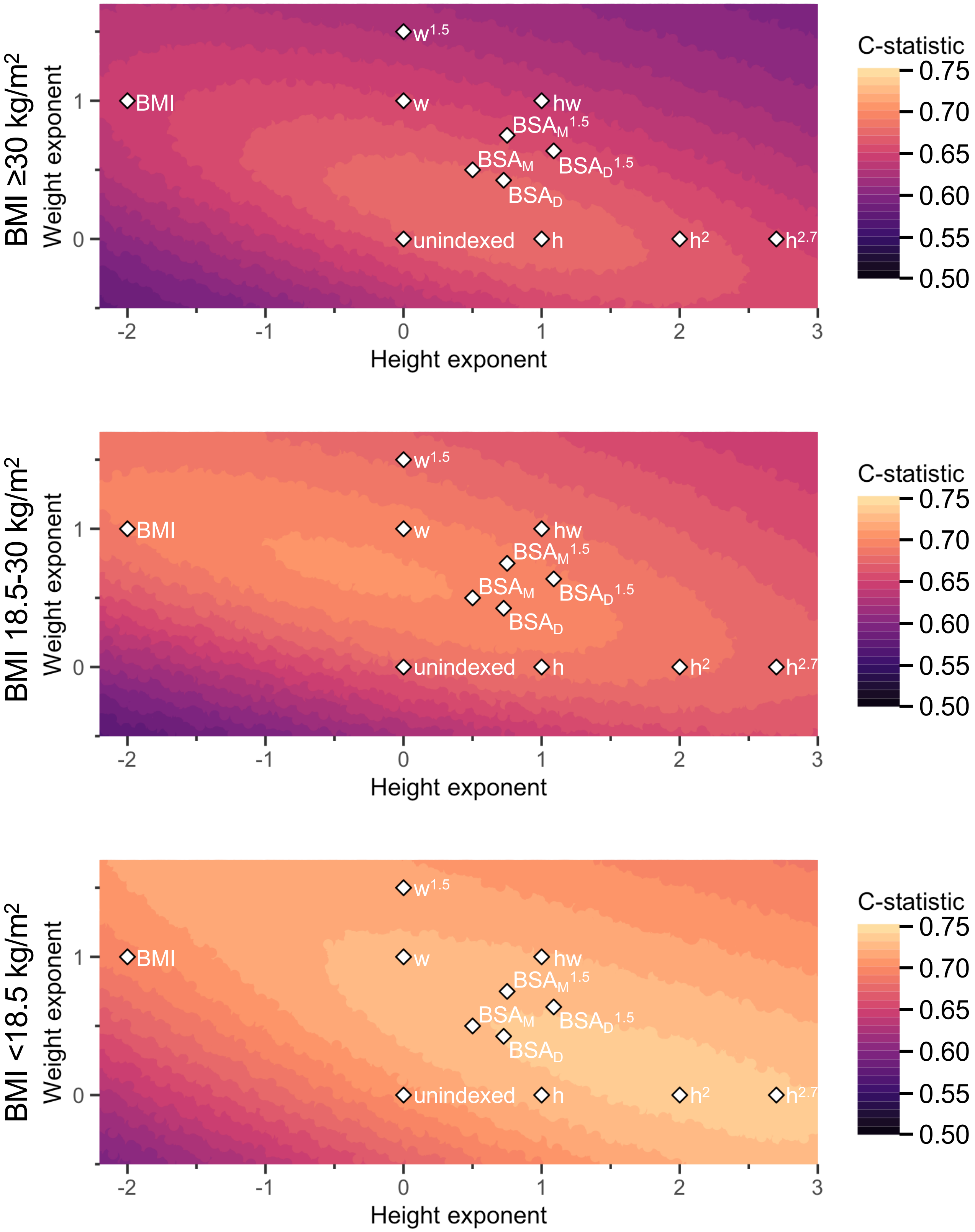


**Supplemental Figure 7.** Average prognostic strength of indexing for body size in LA diameter.

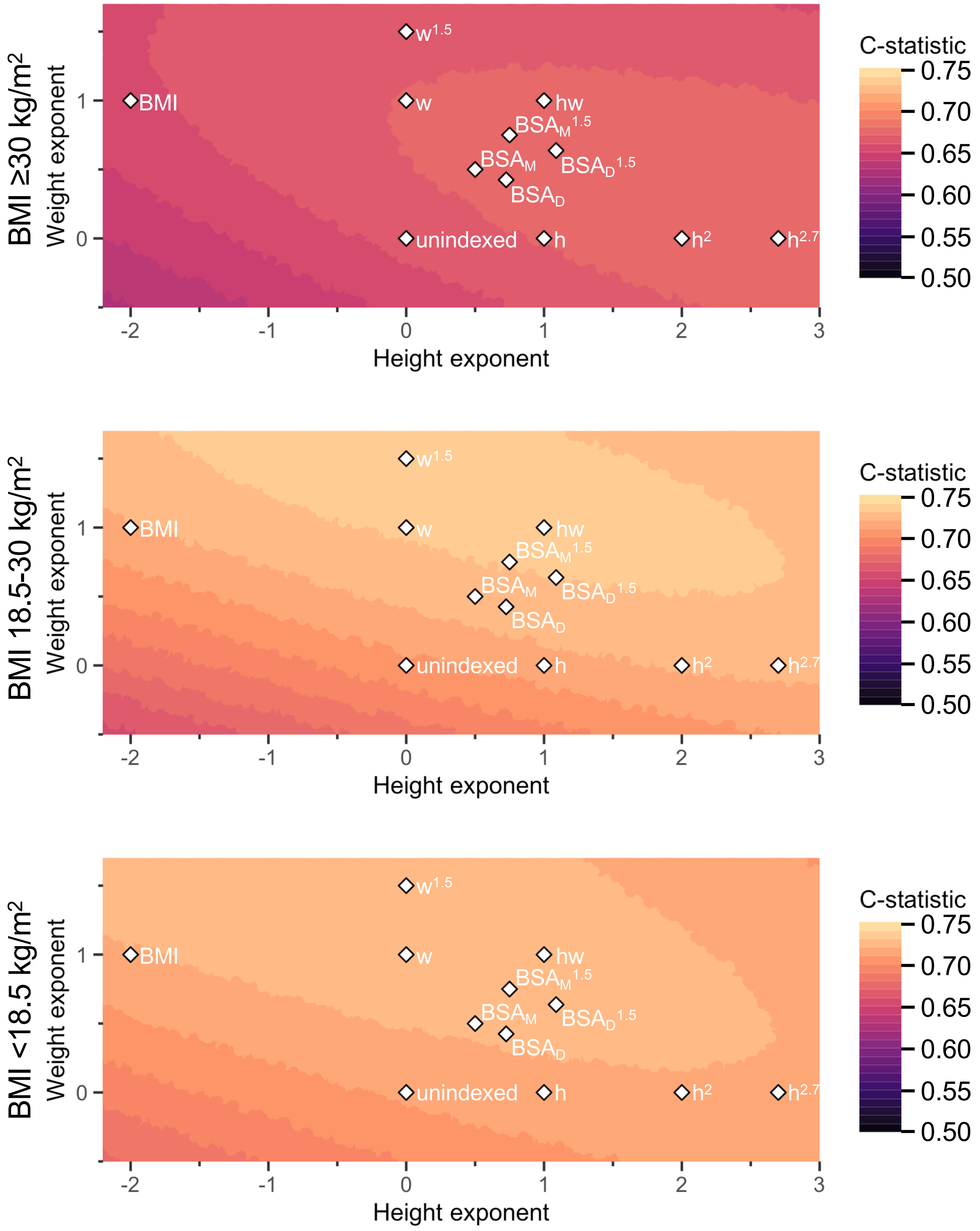


**Supplemental Figure 8.** Average prognostic strength of indexing for body size in LA volume.

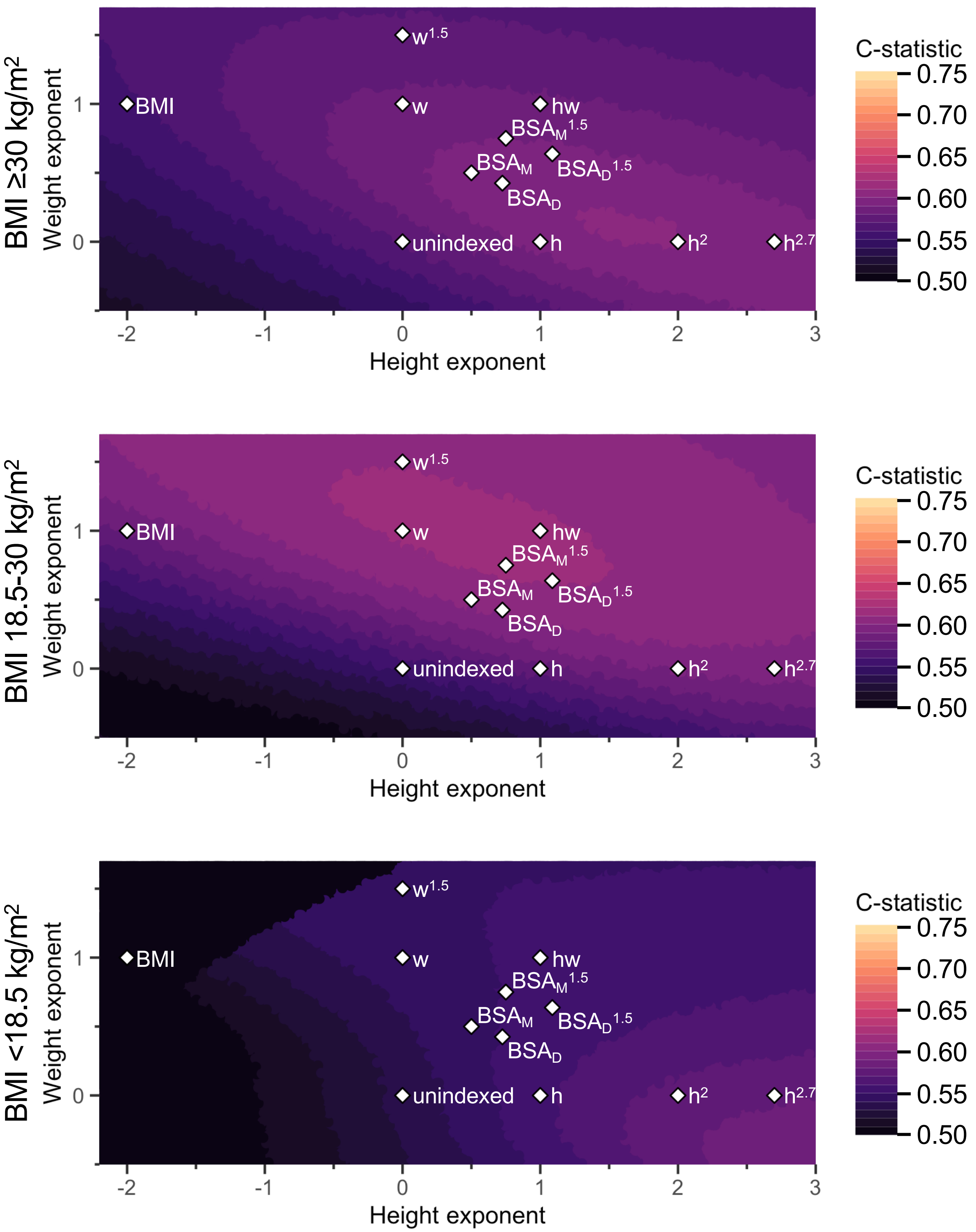


**Supplemental Figure 9.** Average prognostic strength of indexing for body size in LV end-diastolic diameter.

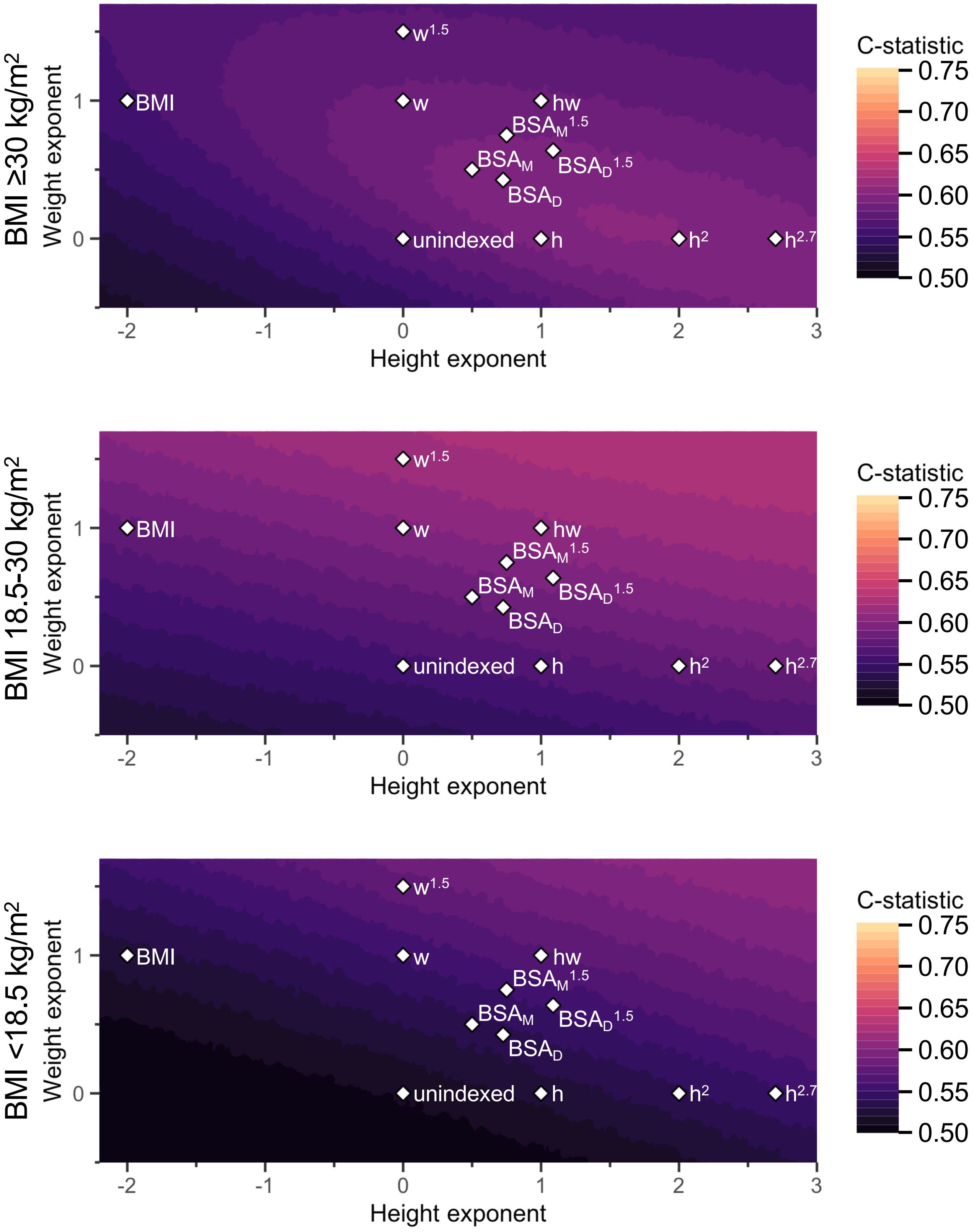


**Supplemental Figure 10.** Average prognostic strength of indexing for body size in LV end-diastolic volume.

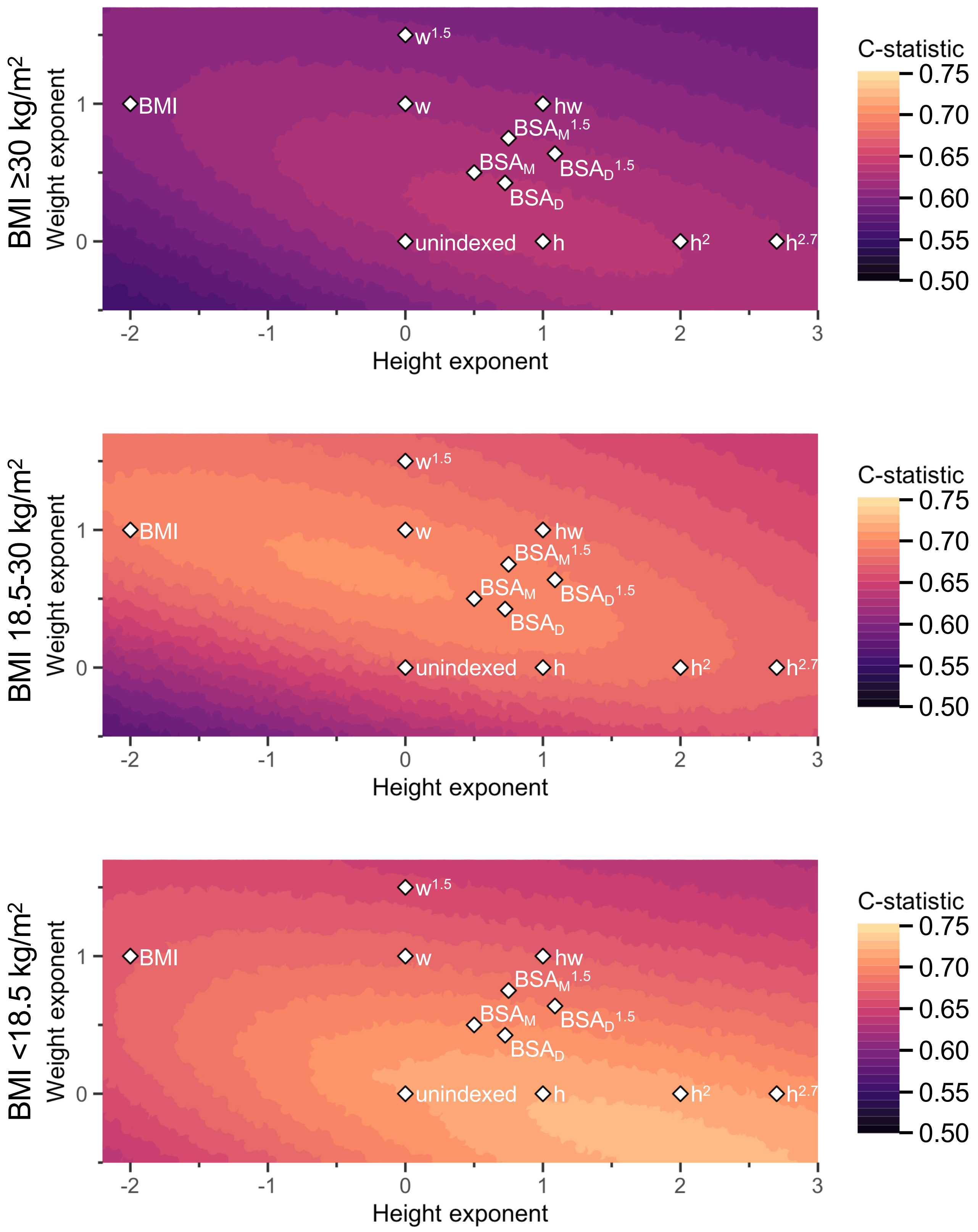


**Supplemental Figure 11.** Average prognostic strength of indexing for body size in IVS thickness.

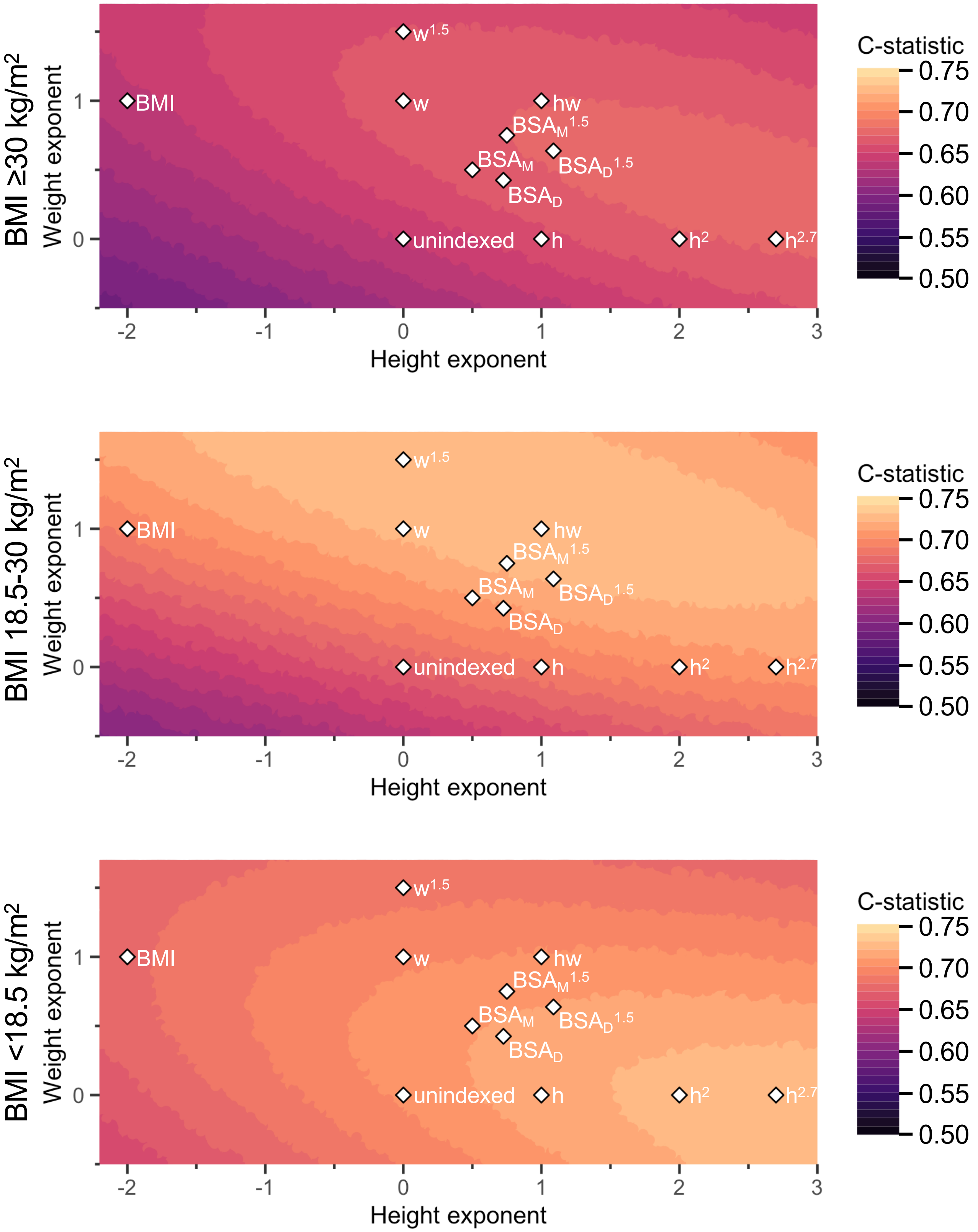


**Supplemental Figure 12.** Average prognostic strength of indexing for body size in LV mass.

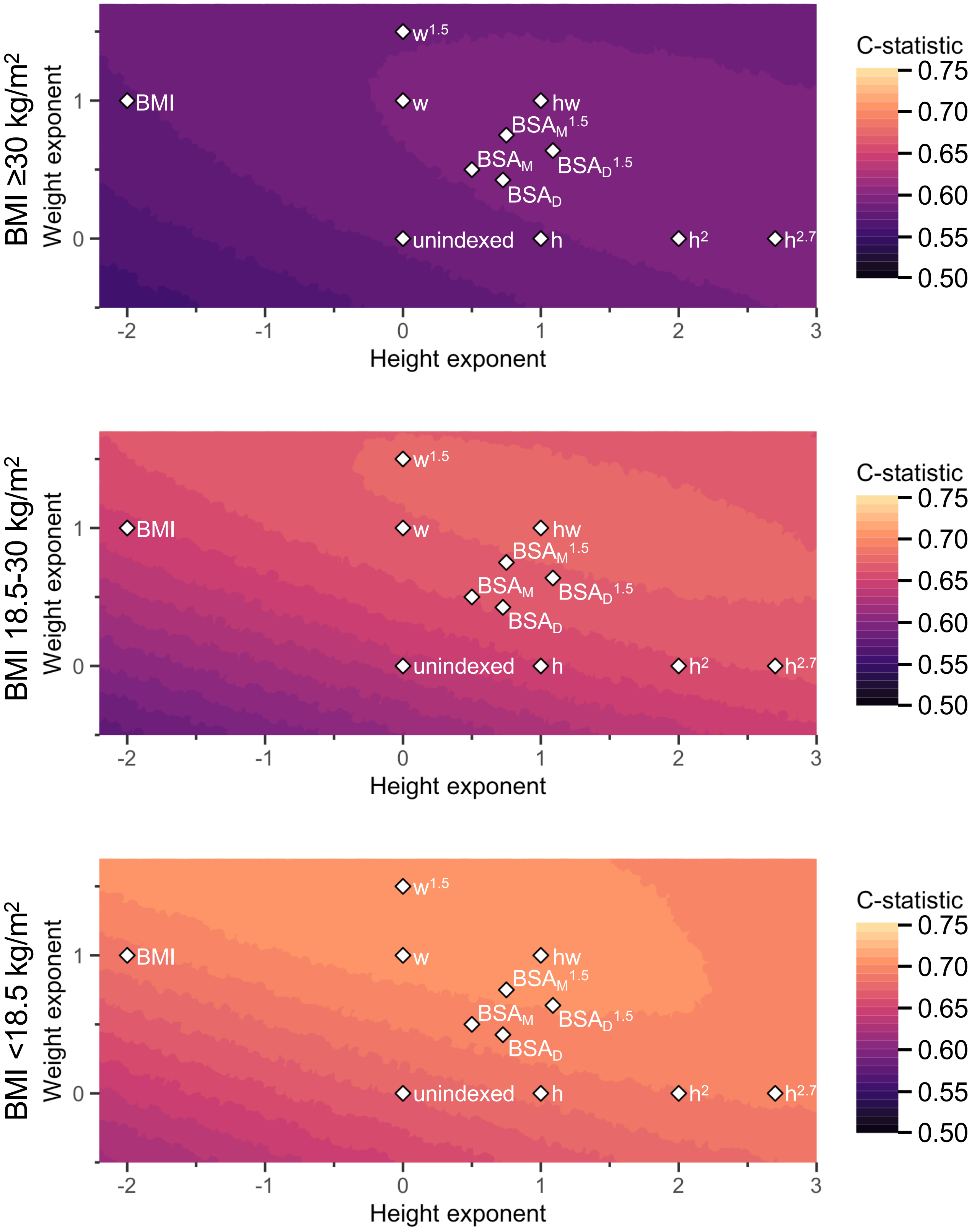


**Supplemental Figure 13.** Average prognostic strength of indexing for body size in RA area.

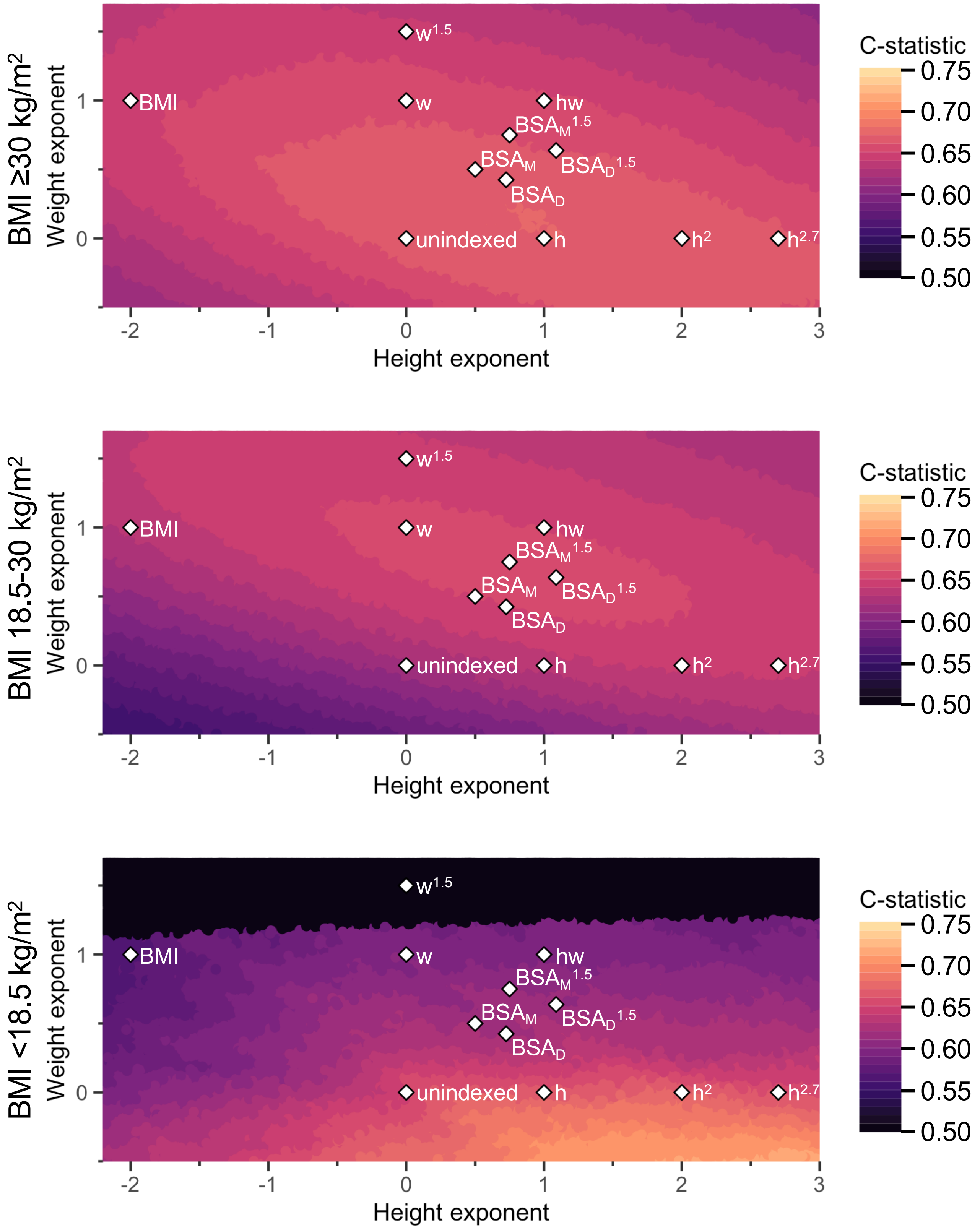


**Supplemental Figure 14.** Average prognostic strength of indexing for body size in RV diameter. Visual artifact present in BMI <18.5 kg/m$^2$ due to small population size (n=93).

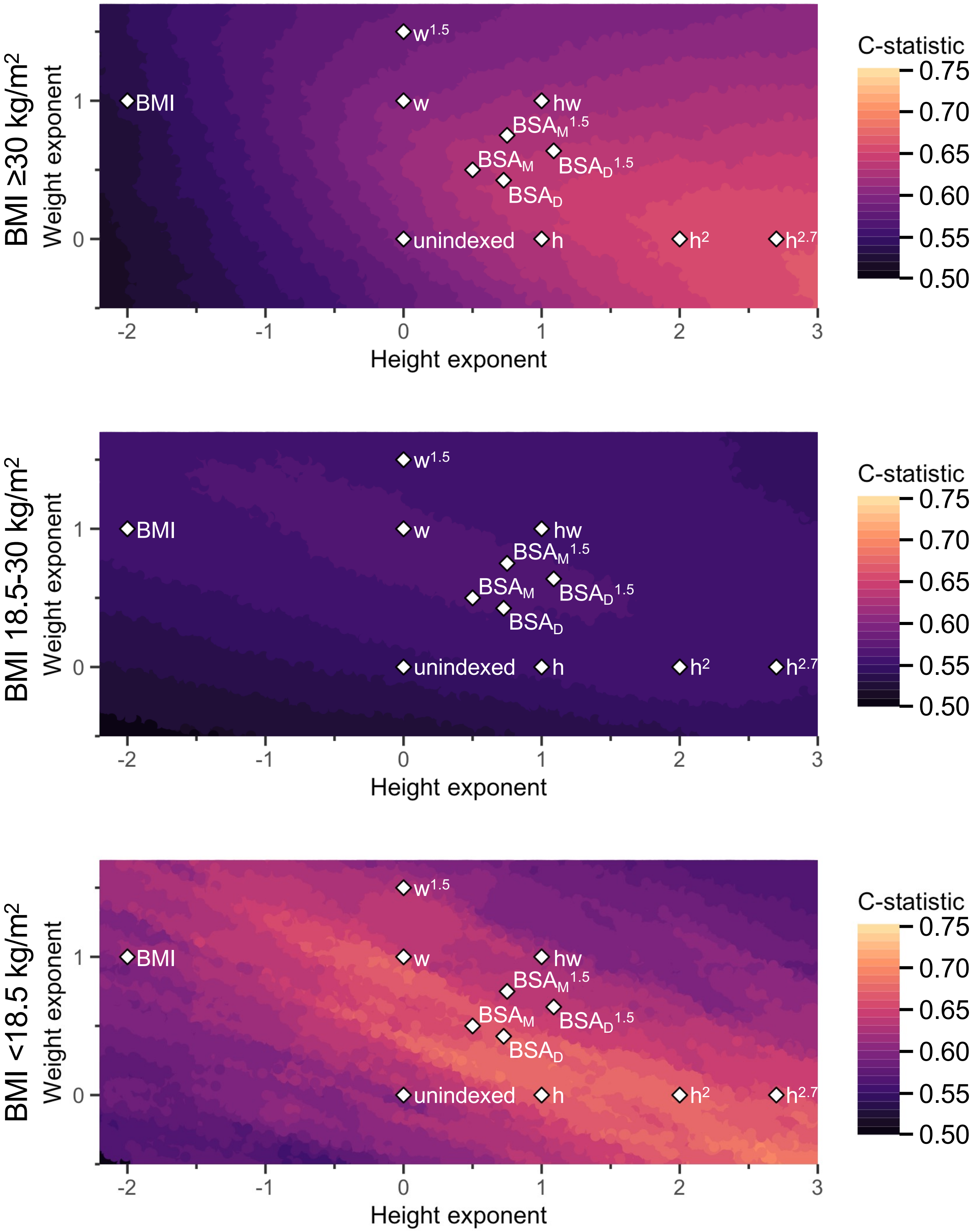


**Supplemental Figure 15.** Average prognostic strength of indexing for body size in RVOT diameter. Visual artifact present in BMI <18.5 kg/m2 due to small population size (n=38).
